# A multicenter review of histopathology of variants in the context of upper urinary tract urothelial carcinoma and their impact on clinical outcomes

**DOI:** 10.1101/2023.06.19.23291581

**Authors:** N Giudici, A Schoch, V Genitsch, JA Rodriguez-Calero, GN Thalmann, R Seiler

## Abstract

**Introduction:** Similar to bladder cancer, about one third of upper tract urothelial carcinoma (UTUC) present variant histology (VH). We aim to evaluate the incidence, clinical characteristics and the impact on outcomes of VH in UTUC.

**Methods:** We consecutively enrolled 77 patients from 2009-2022 treated with radical surgery for UTUC from a secondary and a tertiary referral center. A central pathology review of all specimens was performed by one independent uropathologist for each center. We compared pure UTUC and UTUC with VH and the accuracy of endoscopic biopsy. Descriptive and comparative analysis were used to assess association with clinical characteristics and the Kaplan-Meier estimator to compare outcomes.

**Results:** Median follow-up after surgery was 51 months. VH was present in 21/77 (28%) patients and 4/21 (19%) patients had multiple variants. The most frequent VH was squamous 12/21 (57%), followed by glandular 6/21 (29%) and micropapillary 3/21 (14%). Small cell neuroendocrine bladder carcinoma was present in two patients. Nested variant was found in one patient.

Muscle invasive tumor (≥pT2) was present in 29/56 (52%) patients with pure UTUC and in 18/21 (86%) patients with VH (p <0.05). Presence of carcinoma in situ was seen in 14/56 (25%) patients with pure UTUC and in 15/21 (71%) with VH (p <0.05). Cumulative 8/56 (14%) with pure UTUC had a non-intravesical recurrence (6 patients with local and 2 distant recurrence) compared to 8/21 (38%) (3 local, 3 nodal, 2 distant) in the subgroup with VH (p <0.05). Opposite effect was noted for bladder recurrence: 60% for pure UTUC vs. 29% for tumors with VH (p <0.05). Review of preoperative endoscopic biopsy did not show the presence of VH in any patients. Differences in outcomes did not reach significance: 3yr-OS 63% vs 42% (p 0.28) and 3yr-CSS 77% vs. 50% (p 0.7).

**Conclusion:** Almost a third of UTUC present VH. Presence of VH is related to more aggressive tumor characteristics and associated with unfavorable outcomes. Due to a higher rate of extravesical recurrences in UTUC with VH, Follow-up controls should include cross sectional imaging and cystoscopy.

## Introduction

Urothelial carcinomas are the 6th most common cancers in developed countries (Ferlay et al., 2018)(Siegel et al., 2022). Upper tract urothelial carcinoma (UTUC), accounts for 5-10% of all urothelial carcinoma and pyelocaliceal location of UTUC at diagnosis is twice more common than in the ureter.(Green et al., 2013) The most common symptom in patients with UTUC is haematuria (70-80%) followed by local pain (20-30%) (Inman et al., 2009)^,(Qi et al., 2018; Yeh et al., 2015)^, whilst approximately half of the patients are asymptomatic at presentation (Raman et al., 2011).

Diagnosis of primary UTUC is challenging and based on mainly invasive procedures. Bladder urinary cytology has a relatively low sensitivity for the diagnosis of UTUC and has no role in the diagnosis of UTUC. Invasive procedures such as selective cytology from the UUT show a sensitivity of 70-75% for high grade UTUC (Messer et al., 2011).

In bladder cancer (BC) variant histology (VH) can be observed in about one third of the patients and is an established unfavorable prognostic factor with direct clinical consequences in the oncological and surgical management of the patients. In general a slightly lower incidence of VH (approximately 15-20%) is reported in UTUC (Humphrey et al., 2016)(Mori et al., 2020)(“Effect of Concomitant Variant Histology on the Prognosis of Patients with Upper Urinary Tract Urothelial Carcinoma after Radical Nephroureterectomy,” 2015; Mori et al., 2020). Likely due to the relatively low incidence of UTUC, only few studies have addressed prevalence and prognostic significance of VH in UTUC.

In this study we evaluate the incidence, the clinical characteristics and the impact on outcomes of patients with variant histology of UTUC who underwent radical surgery.

Furthermore we evaluated the performance of preoperative endoscopic biopsy to predict the presence of VH in UTUC.

## 1. Material and methods

### Study Population

We consecutively enrolled 77 patients from 2009-2022 treated with radical nephroureterectomy (RNU) or segmental ureterectomy (SU) in a curative intent with confirmed UTUC from a secondary and from a tertiary referral center. Patients with UTUC who underwent endoscopic treatments or with no tumor in the pathology report were excluded. Preoperative standard diagnostic work-up was performed according to the guidelines for each patient with imaging with computed tomography or magnetic resonance, cystoscopy and selective cytology with or without ureteroscopic biopsy of the tumor in the upper urinary tract (UUT). Surgery was performed by various surgeons with standard lymph node dissection. The decision between SU and RNU was made for each individual patient depending on the characteristics of the tumor, its location and the baseline renal function. A subcohort of patients (n=12) with preoperative endoscopic tumor biopsy was further reviewed to assess the presence of VH in the biopsy specimen.

Each patient included in the cohort signed a declaration of consent of the University Hospital of Bern and of the Hospital Center of Biel for the use of biological material and health-related data for medical research was signed by each patient.

### Pathologic and cytological analysis

A pathology review of all specimens was performed by two independent genitourinary pathologists and classified according to the histological classification published by WHO and the International Society of Urological Pathology in 2016 (Humphrey et al., 2016). All cytologies performed preoperatively were initially evaluated by genitourinary pathologists. Cytology was assessed as either high-grade, atypia/suspicious/low-grade or negative according to the Paris System for reporting urinary cytology (Barkan et al., 2016).

### Statistical analysis

For both cohorts we used descriptive analysis. The p-value were calculated using chi-squared test and Fisher’s Exact Test for categorical variables. Continuous variables were compared using Mann-Whitney U test. Outcomes were calculated using the Kaplan–Meier survival analysis and a p value of less than 0.05 was considered significant. Statistical analysis was performed using IBM SPSS® v. 25 statistical software (SPSS Inc., Chicago, IL, USA).

## 2. Results

### 3.1 Baseline characteristic (Table 1)

In total 77 patients were included. Median age (23 females, 54 males) at surgery was 68 years. History of tobacco consumption was positive in 33/77 (43%) patients, 24/77 (31%) never smoked while the rest was not reported. UTUC were located pelvic-calyceal, ureteral or multifocal in 34/77 (44%), 21/77 (27%) or 22/77 (29%) patients, respectively. At diagnosis, hydronephrosis was present in 52/77 (68%) of the patients. Ureteroscopic biopsy was performed in 46/77 (60%) of the patients and 29/46 (63%) presented high-grade UC, while the rest was low-grade or negative. Local cytology (barbotage and non-barbotage) was analysed in 69/77 (90%) of cases of which high-grade, atypia/suspicious/low-grade and negative, were respectively present in 23/69 (33%), 23/69 (33%) and 23/69 (33%) of the cases. All patients underwent radical surgery: RNU in 55/77(71%) and SU in 22/77 (29%) patients. Muscle-invasive UTUC was present post operatively in 48/77 (62%) patients. High- and low-grade UTUC were found in 65/77 (84%) and 12/77 (16%), respectively. Concomitant carcinoma in situ was found in 29/77 (38%) of the patients. Positive nodal status (pN+) was present in 15/77 (19%) cases. Two patients had positive surgical margins (R1). Median follow-up was 51 (range 1-116) months. It was performed in accordance with the European guidelines (EAU) (Rouprêt et al., 2021) with endoscopic, cytologic and radiologic controls depending on pathological analysis of the tumor.

The total extra-vesical recurrence rate was 25% (63% local, 16% nodal and 21% distant). Bladder recurrence was present in 40/77 (52%) patients after a median of 7 months (range 2-53). The 3-yr Overall survival was 57% and the 3-yr Cancer specific survival 72%.

### 3.2 Prevalence of variant histology (Table 2. Fig 1.)

After central pathological review, 21/77 (27%) UTUC presented variant histology. Of all variants, squamous variant was most frequent (12/21 (57%), including 4 UTUC with multiple variant histologies). In total 5-20% of the examined tumor area showed variant morphology. Glandular variant was present in 6/21 (29%) and observed in 5-30% of the examined tumor area.

Micropapillary histologic variant was identified in 3/21 (14%) of UTUC with variant histology and present in 20-70% of examined tumor area. One patient presented a nested variant in an area of 10% of the UTUC. Primary neuroendocrine carcinoma, was present in two patients both of which had 10% concomitant mucinous differentiation.

### 3.2 Comparison of pure UTUC and UTUC with VH

Table 1 shows the association of pure UC and VH and clinicopathological characteristics.

#### Association of VH with clinicopathologic tumor characteristics

Overall, variant histology was associated with unfavorable tumor characteristics. Muscle invasive tumor stage (≥pT2) was more frequent in patients with VH (18/21, 86%) when compared to patients with pure UC (29/56, 52%), (p value 0.008, Fisher test). Concomitant carcinoma in situ was present in 15/21, 71% and 14/56, 25% with VH and pure UTUC, respectively (p value 0.0002, chi-square test). More patients with VH had positive lymph nodes (33%) when compared to patients with pure UTUC (14 %), however, this did not reach significance (p value 0.148, chi-square test).

Other clinicopathologic parameters such as age, gender, tobacco consumption, tumor location, tumor multifocality, preoperative hydronephrosis, cytology and positive surgical margins were not statistically different between the two subgroups.

#### Prognostic significance of presence of variant histology

In line with the association of unfavorable tumor characteristics, patients with UTUC with VH had an unfavorable prognosis. The rate of extravesical recurrence was higher in patients with variant histology when compared to patients with pure UTUC (38% vs. 14%, p value 0.02, chi-square test). In contrast, recurrence in the bladder was more frequent in pure UTUC when compared to VH (60% vs. 29%; p value 0.0119, chi-square test). Consequently, cause of death due to extravesical recurrence was more frequent in UTUC with VH (5/21, 23%) when compared to pure UTUC (3/56, 5%).

Patients with variant histology had shorter overall and cancer-specific survival when compared to patients with pure UTUC. However this difference did not reach significance (3yr-OS 42% vs 63%, p = 0.28; 3yr-CSS 50% vs. 77%, p = 0.7, Figure xxx (KM curve)).

#### Performance of preoperative endoscopic biopsy in detecting variant histology

Next we investigated whether VH can be detected in the endoscopic biopsy specimen. Therefore, we performed histological review of those patients with VH and preoperative ureteroscopic tumor biopsy. In a total of 8 patients with VH ureteroscopic tumor biopsy was performed. Although all patients presented high-grade UC at biopsy and/or positive cytology, none of the reviewed endoscopic biopsies could detect the presence of variants before radical surgery was performed. This observation reflects the technical limitations of endoscopic diagnostics in the UUT and the often small specimens harvested to confirm diagnosis of UTUC.

## Discussion

This study investigated the incidence, clinical characteristics and impact on outcomes of VH in UTUC. In line with previous series, the results indicate an association of VH in UTUC with more aggressive tumor characteristics, higher recurrence rates and a trend towards worse outcomes (Mori et al., 2020; Yu et al., 2019; Zamboni et al., 2019). In contrast to previous studies and to the best of our knowledge, this is the first study in which a central pathological review and investigation of the accuracy of endoscopic biopsy has been performed.

In our study the incidence of VH is virtually the same when compared to the literature. Around 30% of the patients undergoing radical surgery for UTUC present VH (Mori et al., 2020; Yu et al., 2019)(Yu et al., 2019; Zamboni et al., 2019). This incidence is similar when compared to VH of BC (6-33%) (Veskimäe et al., 2019)(Humphrey et al., 2016) Our central pathological review revealed a significant association of presence of concomitant carcinoma in situ (CIS) in UTUC with VH. This is in contrast to other retrospective series that did not find this association between presence of VH and CIS. (Zamboni et al., 2019). This observation is particularly relevant because CIS is known to be associated with more aggressive tumor features (Wheat et al., 2012) This difference from our study to previous findings, is explained by the pathological review. Presence of CIS was initially only reported in 29 patients, whereas the pathological review revealed CIS in 40 patients.

VH in bladder cancer has been associated with more aggressive tumor characteristics and worse outcomes (“Impact of Histological Variants on Oncological Outcomes of Patients with Urothelial Carcinoma of the Bladder Treated with Radical Cystectomy,” 2013)(Veskimäe et al., 2019)(Royce et al., 2018). Consequently, VH in bladder cancer affects treatment recommendations. Bladder-sparing approaches in T1 bladder cancer with VH are associated with increased risk of tumor progression and radical cystectomy is recommended (Sanguedolce et al., 2021)(Babjuk et al., 2022). Similarly, UTUC with VH is considered as “high-risk” and kidney-sparing surgery or endoscopic treatments is not recommended (Rouprê t et al., 2021). Our study supports these current recommendations in the EAU guidelines. However, endoscopic biopsy misses VH in most UTUC and therefore, this recommendation can only affect adjuvant treatments. Whether treatment intensification by adjuvant chemotherapy should be offered in UTUC with VH needs further investigation.

Patterns of recurrences after treatment of UTUC affects survival and are important to stratify follow-up. Patients treated for UTUC can present intravesical and extravesical recurrences.. However, in the literature this differentiation is often omitted (Mori et al., 2020; Yu et al., 2019; Zamboni et al., 2019). Therefore, we separately analyzed intravesical and extravesical recurrence rates in patients with or without VH. UTUC without VH showed more frequent intravesical recurrence, while in UTUC with VH extravesical recurrence was more often. Extravesical recurrences have a higher effect on survival and require salvage or palliative therapies. In our cohort, 23% of patients with VH died due to extravesical recurrence. This rate is much higher compared to patients without VH (5%). Consequently, follow-up protocols for patients with UTUC and VH should include cross sectional imaging given the increased risk of extravesical recurrence and subsequent death. Adherence to the follow-up schedule should be considered mandatory for these patients.

The results of this study should be interpreted in the context of its limitations. Despite its retrospective nature, a standardized follow-up protocol has been performed. The sample size is limited, however, to the best of our knowledge this is the first cohort in which a central pathological review has been performed. We excluded UTUC treated with endoscopic techniques which may select cases with more aggressive and advanced UTUC. On the other hand, without this exclusion a comparison of the representativeness of endoscopic biopsy for VH with the final surgical specimen would not be possible.

## 3. Conclusion

The central pathological review in our study identified variant histology in a third of UTUC and in these patients a high rate of concomitant CIS (71%). Presence of VH is related to more aggressive tumor characteristics and associated with unfavorable outcomes. Due to a higher rate of extravesical recurrences in UTUC with VH, follow-up controls should include cross sectional imaging and cystoscopy.

## Data Availability

All data produced in the present work are contained in the manuscript

## Bibliography

Babjuk, M., Burger, M., Capoun, O., Cohen, D., Compérat, E. M., Dominguez Escrig, J. L., Gontero, P., Liedberg, F., Masson-Lecomte, A., Mostafid, A. H., Palou, J., van Rhijn, B. W. G., Rouprêt, M., Shariat, S. F., Seisen, T., Soukup, V., & Sylvester, R. J. (2022). European Association of Urology Guidelines on Non-muscle-invasive Bladder Cancer (Ta, T1, and Carcinoma in Situ). European Urology, 81(1), 75–94.

Barkan, G. A., Wojcik, E. M., Nayar, R., Savic-Prince, S., Quek, M. L., Kurtycz, D. F. I., & Rosenthal, D. L. (2016). The Paris System for Reporting Urinary Cytology: the quest to develop a standardized terminology. Jo 23291581v1urnal of the American Society of Cytopathology, 5(3), 177–188.

Effect of concomitant variant histology on the prognosis of patients with upper urinary tract urothelial carcinoma after radical nephroureterectomy. (2015). Urologic Oncology: Seminars and Original Investigations, 33(5), 204.e9–e204.e16.

Ferlay, J., Colombet, M., Soerjomataram, I., Dyba, T., Randi, G., Bettio, M., Gavin, A., Visser, O., & Bray, F. (2018). Cancer incidence and mortality patterns in Europe: Estimates for 40 countries and 25 major cancers in 2018. European Journal of Cancer, 103, 356–387.

Green, D. A., Rink, M., Xylinas, E., Matin, S. F., Stenzl, A., Roupret, M., Karakiewicz, P. I., Scherr, D. S., & Shariat, S. F. (2013). Urothelial carcinoma of the bladder and the upper tract: disparate twins. The Journal of Urology, 189(4), 1214–1221.

Humphrey, P. A., Moch, H., Cubilla, A. L., Ulbright, T. M., & Reuter, V. E. (2016). The 2016 WHO Classification of Tumours of the Urinary System and Male Genital Organs-Part B: Prostate and Bladder Tumours. European Urology, 70(1), 106–119.

Impact of histological variants on oncological outcomes of patients with urothelial carcinoma of the bladder treated with radical cystectomy. (2013). European Journal of Cancer, 49(8), 1889–1897.

Inman, B. A., Tran, V.-T., Fradet, Y., & Lacombe, L. (2009). Carcinoma of the upper urinary tract: predictors of survival and competing causes of mortality. Cancer, 115(13), 2853–2862.

Messer, J., Shariat, S. F., Brien, J. C., Herman, M. P., Ng, C. K., Scherr, D. S., Scoll, B., Uzzo, R. G., Wille, M., Eggener, S. E., Steinberg, G., Terrell, J. D., Lucas, S. M., Lotan, Y., Boorjian, S. A., & Raman, J. D. (2011). Urinary cytology has a poor performance for predicting invasive or high-grade upper-tract urothelial carcinoma. BJU International, 108(5), 701–705.

Mori, K., Janisch, F., Parizi, M. K., Mostafaei, H., Lysenko, I., Kimura, S., Enikeev, D. V., Egawa, S., & Shariat, S. F. (2020). Prognostic Value of Variant Histology in Upper Tract Urothelial Carcinoma Treated with Nephroureterectomy: A Systematic Review and Meta-Analysis. The Journal of Urology, 203(6), 1075–1084.

Qi, N., Zhang, J., Chen, Y., Wen, R., & Li, H. (2018). Microscopic hematuria predicts lower stage in patients with upper tract urothelial carcinoma. Cancer Management and Research, 10, 4929–4933.

Raman, J. D., Shariat, S. F., Karakiewicz, P. I., Lotan, Y., Sagalowsky, A. I., Roscigno, M., Montorsi, F., Bolenz, C., Weizer, A. Z., Wheat, J. C., Ng, C. K., Scherr, D. S., Remzi, M., Waldert, M., Wood, C. G., Margulis, V., & Upper-Tract Urothelial Carcinoma Collaborative Group. (2011). Does preoperative symptom classification impact prognosis in patients with clinically localized upper-tract urothelial carcinoma managed by radical nephroureterectomy? Urologic Oncology, 29(6), 716–723.

Rouprêt, M., Babjuk, M., Burger, M., Capoun, O., Cohen, D., Compérat, E. M., Cowan, N. C., Dominguez-Escrig, J. L., Gontero, P., Hugh Mostafid, A., Palou, J., Peyronnet, B., Seisen, T., Soukup, V., Sylvester, R. J., van Rhijn, B. W. G., Zigeuner, R., & Shariat, S. F. (2021). European Association of Urology Guidelines on Upper Urinary Tract Urothelial Carcinoma: 2020 Update. European Urology, 79(1), 62–79.

Royce, T. J., Lin, C. C., Gray, P. J., Shipley, W. U., Jemal, A., & Efstathiou, J. A. (2018). Clinical characteristics and outcomes of nonurothelial cell carcinoma of the bladder: Results from the National Cancer Data Base. Urologic Oncology, 36(2), 78.e1–e78.e12.

Sanguedolce, F., Calò, B., Mancini, V., Zanelli, M., Palicelli, A., Zizzo, M., Ascani, S., Carrieri, G., & Cormio, L. (2021). Non-Muscle Invasive Bladder Cancer with Variant Histology: Biological Features and Clinical Implications. Oncology, 99(6), 345–358.

Siegel, R. L., Miller, K. D., Fuchs, H. E., & Jemal, A. (2022). Cancer statistics, 2022. CA: A Cancer Journal for Clinicians, 72(1), 7–33.

Veskimäe, E., Espinos, E. L., Bruins, H. M., Yuan, Y., Sylvester, R., Kamat, A. M., Shariat, S. F., Witjes, J. A., & Compérat, E. M. (2019). What Is the Prognostic and Clinical Importance of Urothelial and Nonurothelial Histological Variants of Bladder Cancer in Predicting Oncological Outcomes in Patients with Muscle-invasive and Metastatic Bladder Cancer? A European Association of Urology Muscle Invasive and Metastatic Bladder Cancer Guidelines Panel Systematic Review. European Urology Oncology, 2(6), 625–642.

Wheat, J. C., Weizer, A. Z., Wolf, J. S., Jr, Lotan, Y., Remzi, M., Margulis, V., Wood, C. G., Montorsi, F., Roscigno, M., Kikuchi, E., Zigeuner, R., Langner, C., Bolenz, C., Koppie, T. M., Raman, J. D., Fernández, M., Karakiewizc, P., Capitanio, U., Bensalah, K., … Shariat, S. F. (2012). Concomitant carcinoma in situ is a feature of aggressive disease in patients with organ confined urothelial carcinoma following radical nephroureterectomy. Urologic Oncology, 30(3), 252–258.

Yeh, H.-C., Jan, H.-C., Wu, W.-J., Li, C.-C., Li, W.-M., Ke, H.-L., Huang, S.-P., Liu, C.-C., Lee, Y.-C., Yang, S.-F., Liang, P.-I., & Huang, C.-N. (2015). Concurrent Preoperative Presence of Hydronephrosis and Flank Pain Independently Predicts Worse Outcome of Upper Tract Urothelial Carcinoma. PloS One, 10(10), e0139624.

Yu, J., Li, G., Wang, A., Luo, Q., Liu, Z., Niu, Y., & Mei, Y. (2019). Impact of squamous differentiation on intravesical recurrence and prognosis of patients with upper tract urothelial carcinoma. Annals of Translational Medicine, 7(16), 377.

Zamboni, S., Foerster, B., Abufaraj, M., Seisen, T., Roupret, M., Colin, P., De la Taille, A., Di Bona, C., Peyronnet, B., Bensalah, K., Herout, R., Wirth, M. P., Novotny, V., Soria, F., Chlosta, P., Antonelli, A., Simeone, C., Baumeister, P., Mattei, A., … European Association of Urology - Young Academic Urologists (EAU-YAU), Urothelial carcinoma working group. (2019). Incidence and survival outcomes in patients with upper urinary tract urothelial carcinoma diagnosed with variant histology and treated with nephroureterectomy. BJU International, 124(5), 738–745.

