## Supplemental Table 1. and 2. for "A multicenter review of histopathology of variants in the context of upper urinary tract urothelial carcinoma and their impact on clinical outcomes"

**Table 1. Clinicopathological characteristics of the UTUC cohort**

| | UTUC cohort (n=77) | | | |
|---|---|---|---|---|
| | All patients | pure urothelial histology (n=56) | variant histology (n=21) | *p value* |
| **Patient data** | | | | |
| Age (median, range) at diagnosis [years] | 68 (44-90) | 68 (44-86) | 71 (46-90) | 0.85 |
| Female/male (n) | 23/54 | 17/39 | 6/15 | 1 |
| History of tobacco smoking/never smoked/not reported (n) | 33/24/14 | 28/16/12 | 11/8/2 | 0.78 |
| **Diagnostic data** | | | | |
| Tumor location pelvic-calyceal/ureter | 34/21 | 24/18 | 10/3 | 0.32 |
| Multifocality (n) | 22 | 14 | 8 | 0.56 |
| Hydronephrosis (n) | 52 | 38 | 14 | 1 |
| Selective cytology (n) | 69 | 50 | 19 | |
| high-grade/ atypia, suspicious, low-grade/ negative | 23/23/23 | 16/17/17 | 7/6/6 | 0.78 |
| **Surgery data** | | | | |
| Radical Nephroureterectomy/segmental ureterectomy | 55/22 | 36/19 | 18/3 | |
| <pT2/≥pT2 (n) | 29/48 | 27/29 | 3/18 | **0.01** |
| pN+ / with extracapsular extension (ECE) (n) | 15/2 | 8/1 | 7/1 | for pN+:0.10<br>for ECE: 0.48 |

| | | | | |
|---|---|---|---|---|
| Concomitant Cis (n) | 29 | 14 | 15 | **0.00** |
| High-grade/Low-grade | 65/12 | 45/11 | 20/1 | 0.2 |
| PSM (n) | 2 | 1 | 1 | 0.48 |
| **Chemotherapy** | | | | |
| neoadjuvant therapy (n) | 2 | 1 | 1 | 0.48 |
| **Follow-up and recurrence (n)** | | | | |
| Follow-up (reverse KM)(median, range) [months] | 51 (1-116) | | | |
| Recurrence total (no bladder) (n) | 19 | 8 (14%) | 8 (38%) | **0.02** |
| local/nodal/ distant (n) | 12/3/4 | 6/0/2 | 3/3/2 | |
| bladder recurrence (n) | 40 (52%) | 34 (60%) | 6 (29%) | **0.01** |
| median time to bladder recurrence [months] | 7 | 7 | 5 | |
| **Outcomes** | | | | |
| 3yr-OS (%) | 57% | 63% | 42% | 0.28 |
| 3yr-CSS (%) | 72% | 77% | 50% | 0.7 |

Table. 2

| Patient N. | Variant (% of the tumor area) |
|---|---|
| 1-8 | Squamous (5-20) |
| 9-12 | Glandular (10-30) |
| 13 | Micropapillary (70) |
| 14 | Nested (10) |

| | |
|---|---|
| 15 | Micropapillary (30), glandular (20), squamous (10) |
| 16 | Glandular (30), squamous (5) |
| 17 | Glandular (5), squamous (10) |
| 18 | Micropapillary (20), squamous (10) |
| 19 | Mucinous after chemotherapy |
| 20-21 | Neuroendocrine (70-90), mucinous (10) |
